# Persistent COVID-19 symptoms minimally impact the development of SARS-CoV-2 specific cellular immunity

**DOI:** 10.1101/2021.01.29.21250771

**Authors:** HengSheng Fang, Adam D. Wegman, Kianna Ripich, Heather Friberg, Jeffrey R. Currier, Stephen J. Thomas, Timothy P. Endy, Adam T. Waickman

## Abstract

SARS-CoV-2 represents an unprecedented public health challenge with many unknowns remaining regarding the factors that impact viral pathogenicity and the development of immunity after infection. While the majority of SARS-CoV-2 infected individuals with mild-to-moderate COVID-19 resolve their infection with few complications, a significant number of individuals experienced prolonged symptoms lasting for weeks after initial diagnosis. Persistent viral infections are commonly accompanied by immunologic dysregulation, especially within the cellular immune compartment. However, it is unclear if persistent mild-to-moderate COVID-19 impacts the development of virus-specific cellular immunity. To this end, we analyzed the development of SARS-CoV-2 specific cellular immunity in convalescent COVID-19 patients who experienced eight days or fewer of COVID-19 symptoms, or symptoms persisting for 18 days or more. We observed that the duration of COVID-19 symptoms minimally impacts the magnitude, antigen specificity, and transcriptional profile of SARS-CoV-2 specific cellular immunity within both the CD4+ and CD8+ T cell compartments. Furthermore, we observed that reactivity against the structural N protein from SARS-CoV-2 in convalescent COVID-19 patients correlates with the amount of reactivity against the seasonal human coronaviruses 229E and NL63. These results provide additional insight into the complex processes that regulate the development of cellular immunity against SARS-CoV-2 and related human coronaviruses.

## INTRODUCTION

SARS-CoV-2 is a recently emerged novel single-stranded RNA virus that was initially identified as the causative agent of a pneumonia outbreak in Wuhan, China in early December, 2019 [1-3]. This initial outbreak has since developed into an unprecedented world-wide pandemic, resulting in an estimated 96 million infections and 2 million deaths as of January 2021. The multi-faceted illness associated with SARS-CoV-2 infection – COVID-19 – is characterized by inflammation of the respiratory tract, fever, musculoskeletal pain, and cough [4-6]. While SARS-CoV-2-specific humoral and cellular immunity is evident in the majority of patients following the resolution of acute infection and appears to persist for at least 6-8 months [7, 8], the role of this adaptive immune response in regulating viral replication and disease pathogenesis remains unclear. Furthermore, little is known about how variations in the complex clinical manifestations of COVID-19 impact the development of SARS-CoV-2 specific immunologic memory.

A notable feature of SARS-CoV-2 infection is that COVID-19 symptoms can persist for weeks or months after initial manifestation even in patients not requiring hospitalization or other medical interventions [9, 10]. This is especially evident in older adults with underlying chronic medical conditions but has been extensively documented in patients across a wide age range [10]. Even in young adults, nearly 20% of patients with confirmed SARS-CoV-2 infection fail to return to full normal daily activities 14-21 days after the onset of COVID-19 symptoms and/or a positive SARS-CoV-2 test in an outpatient setting [11]. Although replication-competent SARS-CoV-2 has been difficult to detect in individuals with protracted COVID-19 symptoms, recovered patients continue to shed detectable SARS-CoV-2 RNA in their upper respiratory tract and in their stool for weeks after initial diagnosis [12-14]. Furthermore, indirect immunologic evidence of SARS-CoV-2 antigen persistence has been observed, most notably reflected in the maturation profile of SARS-CoV-2 specific memory B cells [15].

The presence of persistent viral antigen and/or infection-attendant inflammation is associated with immune dysregulation in many viral infections [16]. This is most prominently manifested within the cellular immune compartment where persistent antigen stimulation and/or inflammatory cytokine exposure can lead to a progressive loss of T cell effector function and suppression of pathogen-specific cellular immunity [17, 18]. While this phenomenon has been well documented in chronic viral diseases such as HBV/HCV and HIV [18], it is currently unclear if the persistence of mild-to-moderate COVID-19 symptoms is associated with the development of a dysfunctional or sub-optimal cellular immune response.

To fill this knowledge gap, we examined the relationship between the duration of COVID-19 symptoms and the magnitude and functional profile of SARS-CoV-2 specific cellular immunity in individuals recently recovered from mild-to-moderate COVID-19. Using an IFN-γ ELISPOT assay, we observed that patients with protracted COVID-19 symptoms exhibited similar levels of SARS-CoV-2 specific cellular immunity overall as individuals who rapidly resolved their symptoms; although prolonged COVID-19 symptoms were associated with slightly elevated responses against SARS-CoV-2 ORF3a and ORF7a. Furthermore, no defect was observed in the magnitude of the SARS-CoV-2 Spike specific CD4+ and CD8+ T cell response in individuals with prolonged COVID-19 symptoms when assessed using flow cytometry, and the transcriptional profile of SARS-CoV-2 specific CD4 T cells was observed not to be impacted by the duration of COVID-19 symptoms. Finally, while significant levels of cellular immunity against the seasonal human coronaviruses 229E and NL63 were observed in all convalescent COVID-19 patients analyzed in the study, the magnitude of this immune response did not correlate with the duration of COVID-19 symptoms. However, a significant negative correlation was observed between patient age and the overall magnitude of 229E/NL63 reactivity across all donors, and the level of 229E and NL63 Spike reactivity did correlate with reactivity against the structural N protein from SARS-CoV-2. These data suggest that prolonged symptomatic COVID-19 does not significantly impact the development of SARS-CoV-2 specific cellular immunity or cellular immunity against related seasonal human coronaviruses in patients with mild/moderate disease, but that prior infection with seasonal human betacoronaviruses may selectively influence the development of N specific SARS-CoV-2 cellular immunity.

## RESULTS

### Convalescent COVID-19 patient selection and characterization

The objective of this study was to determine the impact of COVID-19 symptom duration on the magnitude and functional profile of SARS-CoV-2 specific cellular immunity. To this end, subjects were identified within the SUNY Upstate Convalescent COVID-19 Plasma Donor protocol who experienced a PCR confirmed SARS-CoV-2 infection and from whom PBMC were obtained 14 to 30 days following the resolution of COVID-19 associated symptoms (**Figure 1A**). A total of 84 subjects were identified within the parental protocol who fulfilled these selection criteria, of which 33 were selected for further analysis (**Table 1)**. Within this group of 33 donors, 14 subjects were classified as having a “short” period of COVID-19 associated symptoms (0-8 days), while 19 subjects were classified as having a “long” duration of COVID-19 associated symptoms (18-61 days). No correlation was observed between subject age and the duration of self-reported COVID-19 symptoms (**Figure 1B**), and all subjects were otherwise healthy at the time of PBMC collection.

**Table 1.** Convalescent COVID-19 patient characterization

|  | **All subject (n = 84)** | **Short duration of symptoms (n -14)** | **Long duration of symptoms (n – 19)** |
| --- | --- | --- | --- |
| **Age (mean)** | 49 (20 – 85) | 49 (22-65) | 52 (23-72) |
| **Duration of symptoms (mean)** | 14 days (0-61) | 4.6 days (0-8) | 26.7 days (18-61) |
| **Sex (M/F)** | 33/51 | 7/7 | 3/16 |

**Figure 1.**
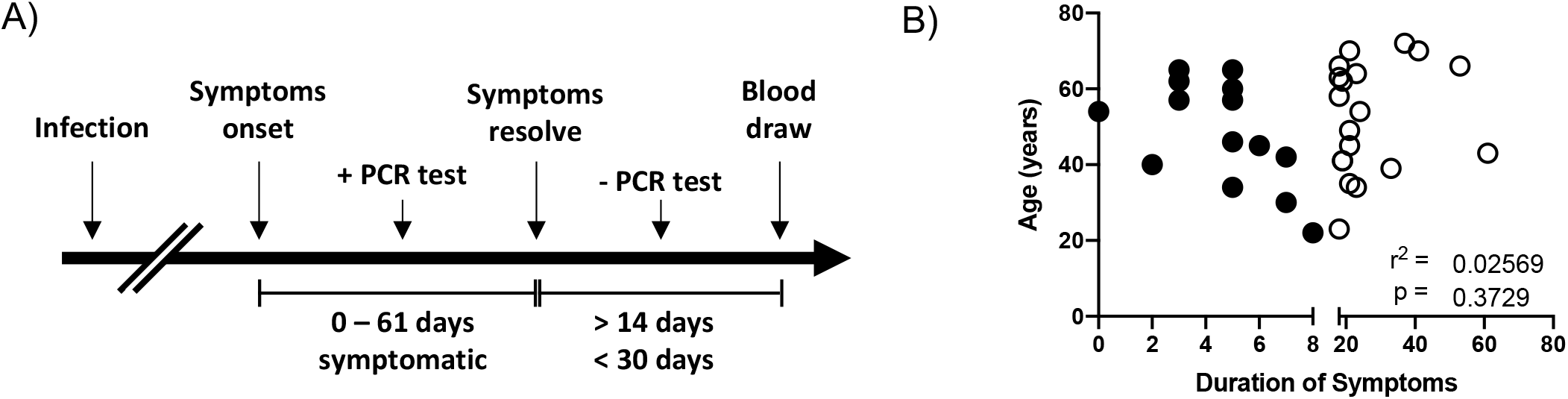
Study design and patient characteristics. **A)** Schematic representation of the subject selection criteria used in this study and the timing of sample collection. **B)** Analysis of the relationship between subject age and the duration of self-reported COVID-19 symptoms in all subjects included in this analysis. Filled circles indicate subjects included in the “short” duration of symptoms group. Empty circles indicate individuals included in the “long” duration of symptoms group. r^2^ and p value calculated by 2-tailed Pearson Correlation test.

### Assessment of SARS-CoV-2 specific cellular immunity stratified by COVID-19 symptom duration

To determine if the magnitude and antigen-specificity of SARS-CoV-2 elicited cellular immunity is impacted by the duration of COVID-19 symptoms, PBMC from the 33 subjects selected above were analyzed using an IFN-γ ELISPOT assay. Overlapping peptide pools spanning the Spike, N, M, ORF3a, and ORF7a proteins from SARS-CoV-2 were used in this analysis (**Supplemental Table 1**). SARS-CoV-2 specific cellular immunity – defined as a subject having more than 50 IFN-γ producing SARS-CoV-2 specific cells per 10^6^ PBMC - was observed in 85.7% (13/14) of subjects classified as having a short duration of COVID-19 symptoms, while 94.7% (18/19) of subjects with a long duration of COVID-19 symptoms exhibited a positive response (**Table 1**). However, no difference in the total SARS-CoV-2 specific cellular immune response was observed between these two groups when stratified by the duration of self-reported COVID-19 symptoms (**Figure 2A, Supplemental Figure 1**). When further stratified by viral antigen, no difference in the level of reactivity against SARS-CoV-2 Spike, N, and M was observed between individuals with either a short or long duration of COVID-19 symptoms (**Figure 2B**). A statistically significant higher level of ORF3a and OFR7a reactivity was observed in individuals with longer periods of COVID-19 symptoms than in individuals with a short period of COVID-19 associated symptoms (**Figure 2B**), but most of these responses fell under the 50 SFC/10^6^ PBMC threshold for positivity (**Table 1**).

**Figure 2.**
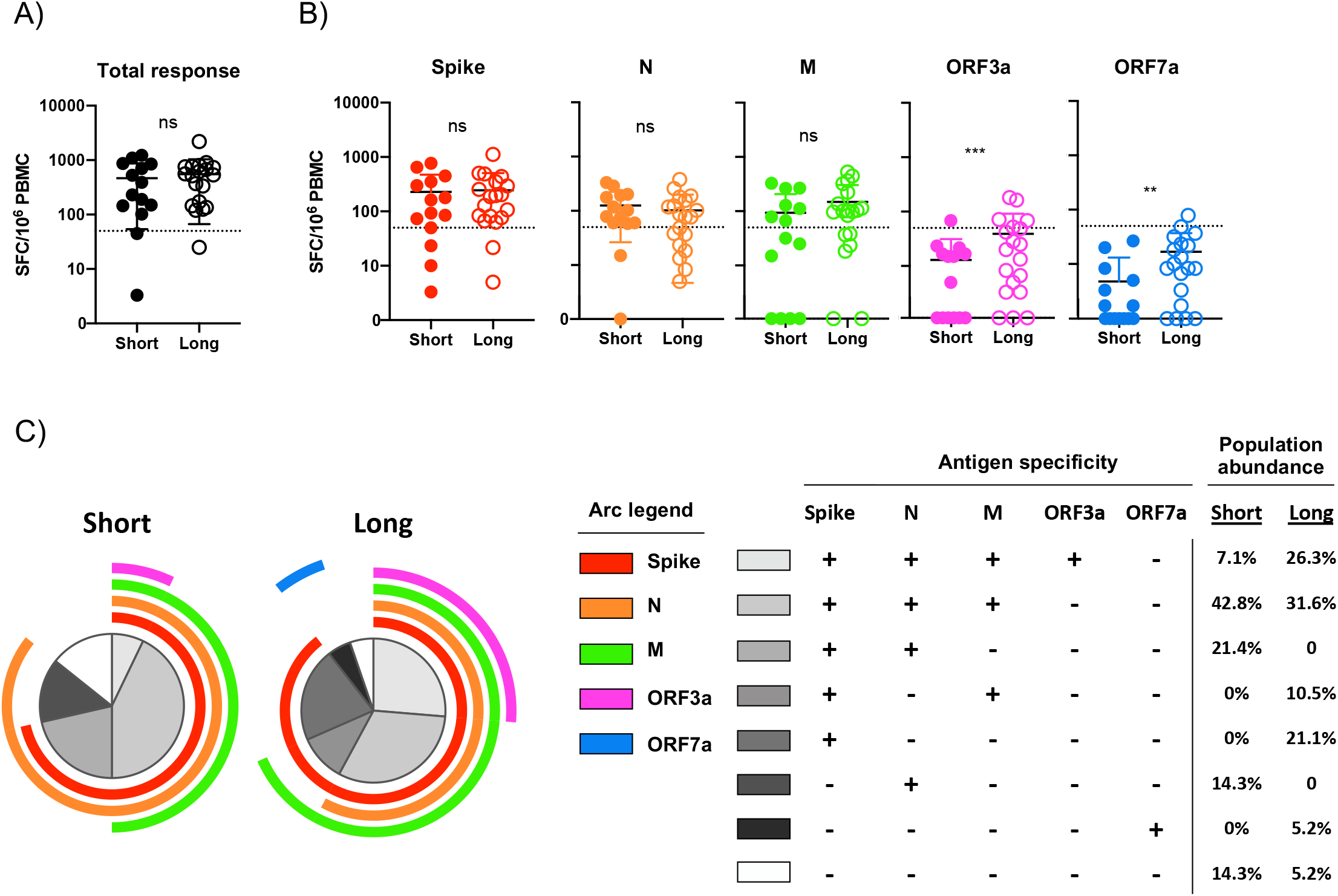
Assessment of SARS-CoV-2 specific cellular immunity by IFN-γ ELISPOT. **A)** Total magnitude of SARS-CoV-2 specific cellular immunity in all study participants as defined by total reactivity against SARS-CoV-2 Spike, N, M, ORF3a and ORF7a antigens. Subjects split by duration of self-reported symptoms. Dashed line indicates a 50 SFC/10^6^ PBMC threshold for a positive response. **B)** Magnitude of SARS-CoV-2 specific cellular immunity separated by major antigen in all study participants. Dashed line indicates a 50 SFC/10^6^ PBMC threshold for a positive response. **C)** Pattern of multi-antigen SARS-CoV-2 reactivity in all study subjects split by duration of self-reported symptoms. Arc color and arc length indicates reactivity against a given SARS-CoV-2 antigen. Internal plot wedge size indicates fraction of individuals with the indicated pattern of antigen reactivity. ^***^ p < 0.001, ^**^ p < 0.01 unpaired 2-tailed T test

To further define the profile of SARS-CoV-2 specific cellular immunity and how it stratifies by COVID-19 symptom duration, we assessed the multi-parametric antigen reactivity pattern captured in our ELISPOT analysis. Most individuals included in this analysis exhibited cellular immunity against two-or-more SARS-CoV-2 antigens, with 71.3% of individuals with a short period of COVID-19 symptoms and 68.4% of individuals that experienced a long period of COVID symptoms exhibiting a multivalent antigen response (**Figure 2C**). The most common multi-antigen reactivity pattern observed in individuals with a short duration of COVID-19 symptoms was a trivalent response against SARS-CoV-2 Spike, N, and M proteins (**Figure 3C**). In contrast, the most common antigen reactivity pattern observed in individuals with a long duration of COVID-19 symptoms was a tetravalent response against SARS-CoV-2 Spike, N, M and ORF3a.

**Figure 3.**
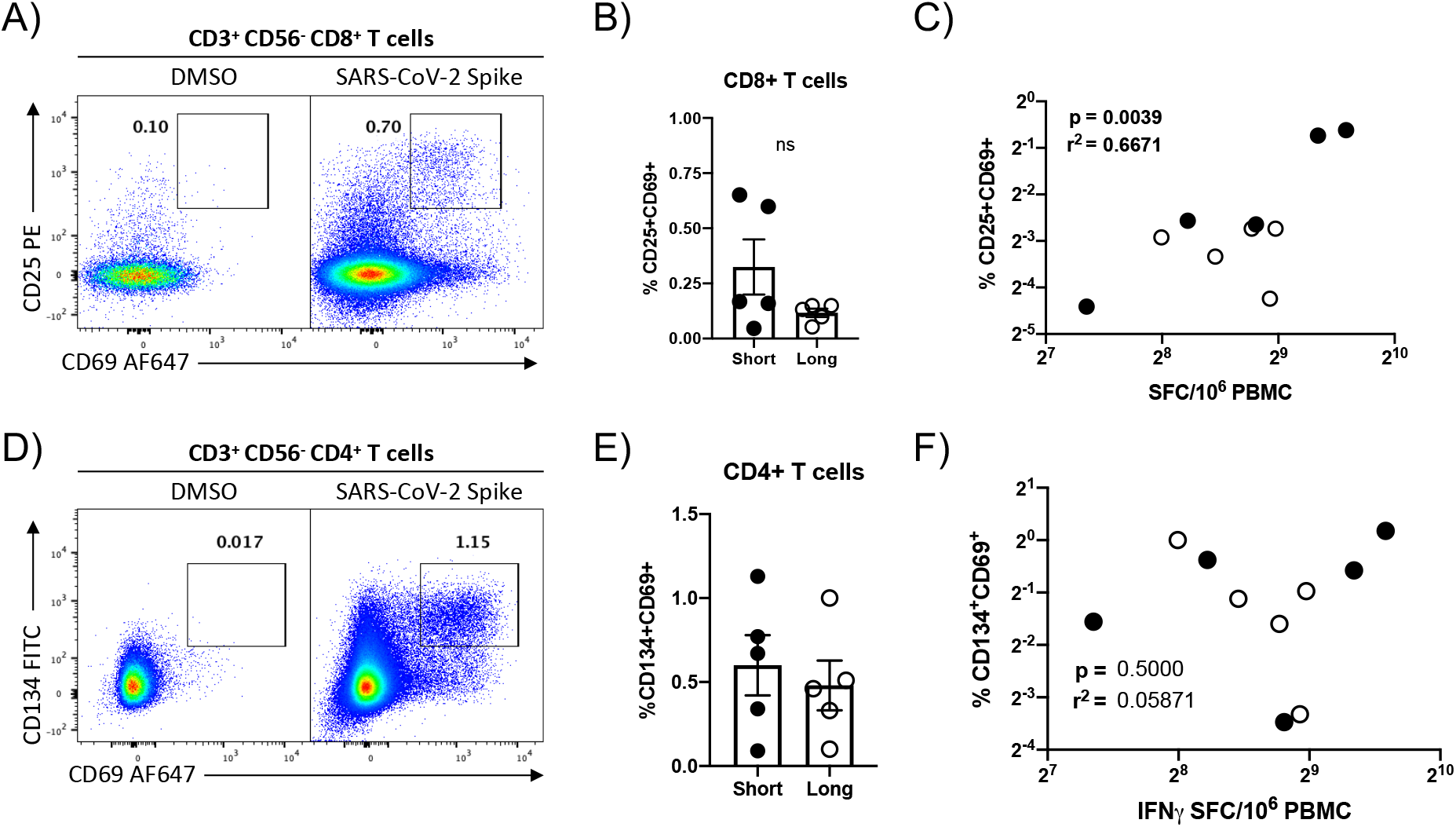
Flow cytometric quantification of SARS-CoV-2 Spike protein specific cellular immunity. **A)** Representative flow cytometry plot demonstrating CD25 and CD69 upregulation in CD8+ T cells following *in vitro* SARS-CoV-2 Spike protein peptide pool stimulation. **B)** Magnitude of SARS-CoV-2 Spike-specific CD8^+^ T cell responsiveness in select study participants split by duration of self-reported duration of COVID-19 symptoms. Plotted values are background subtracted from a total of 10 subjects. **C)** Correlation analysis of SARS-CoV-2 specific cellular immunity as defined by CD8+ flow cytometry and IFN-g ELISPOT. r^2^ and p value calculated by 2-tailed Pearson Correlation test. **D)** Representative flow cytometry plot demonstrating CD134 and CD69 upregulation in CD4+ T cells following SARS-CoV-2 Spike protein peptide pool stimulation. **E)** Magnitude of SARS-CoV-2 Spike-specific CD4+ T cell immunity in select study participants split by duration of self-reported duration of COVID-19 symptoms. Plotted values are background subtracted from a total of 10 subjects. **F)** Correlation analysis of SARS-CoV-2 specific cellular immunity as defined by CD4+ flow cytometry and IFN-g ELISPOT. r^2^ and p value calculated by 2-tailed Pearson Correlation test.

### Flow cytometric assessment of SARS-CoV-2 antigen reactivity

To further define the cellular disposition of the SARS-CoV-2 specific cellular immune response quantified in our ELISPOT analysis, a second aliquot of PBMC from 10 convalescent COVID-19 patients (5 with short symptoms, 5 with prolonged symptoms) that were determined to have high levels of SARS-CoV-2 specific cellular immunity were stimulated with a SARS-CoV-2 Spike protein peptide pool and the level of SARS-CoV-2 specific cellular immunity quantified by flow cytometry. SARS-CoV-2 reactive CD8+ and CD4+ T cells were identified by their respective upregulation of CD25/CD69 or CD134/CD69 following peptide stimulation. A robust population of SARS-CoV-2 reactive CD8+ T cells were identified in the subjects selected for analysis (**Figure 3A, Supplemental Figure 2**), although the abundance of these cells did not significantly differ when stratified by the duration of COVID-19 symptoms (**Figure 3B**). The magnitude of SARS-CoV-2 Spike protein specific cellular immunity measured by IFN-γ ELISPOT correlated well with the CD8+ T cell SARS-CoV-2 specific cellular immunity as quantified by flow cytometry (**Figure 3C**). In addition to this SARS-Cov-2 Spike protein specific CD8+ T cell response, a quantifiably more robust CD4+ T cell response was observed in all donors **(Figure 3A, Supplemental Figure 2)**, with most subjects exhibiting a ∼50% higher frequency of SARS-CoV-2 Spike reactive CD4 T cells than CD8 T cells (**Supplemental Figure 3)**. Again, the overall magnitude of the SARS-CoV-2 specific CD4+ T cell response was not impacted by the duration of COVID-19 symptoms. However, there was a very poor correlation between SARS-CoV-2 Spike protein specific CD4+ T cell response as quantified by flow cytometry and the SARs-CoV-2 Spike reactivity as quantified by IFN-γ ELISPOT, highlighting the differential utility of the two assays.

### Transcriptional characterization of SARS-CoV-2 reactive CD4+ T cells

Persistent antigen simulation is known to result in transcriptional- and functional-dysregulation of pathogen-specific T cells and loss of effector function [19]. While no significant difference in the abundance of SARS-CoV-2 reactive CD4 and CD8 T cells were noted in the convalescent COVID-19 patients included in this study when stratified by symptom duration, we wished to confirm that the functional transcriptional profile of SARS-CoV-2 reactive CD4+ T cells was not negatively impaired in individuals experiencing prolonged COVID-19 symptoms. Therefore, we sorted SARS-CoV-2 Spike protein reactive CD4 T cells from the 10 donors highlighted above and subjected them to transcriptional profiling. An average of 2,322 SARS-CoV-2 reactive CD4+ T cells were isolated from each donor (range 106 – 4,497) and were subjected to mRNA sequencing analysis.

As expected, the cells recovered in this analysis expressed high levels of canonical CD4+ T cell gene products (CD3E, CD4, CD40LG) along with the activation markers used to identify/isolate the cell in the flow cytometry assay (**Figure 4A**). The sorted SARS-CoV-2 specific CD4+ T cells expressed high levels of canonical Th1-assocaited gene products (IFNG, TNF, TBX21), but appreciably lower levels of Th2/Th17 associated transcripts (**Figure 4B**). However, no differentially expressed genes were identified between the samples when segregated by symptom duration, and no appreciable difference was observed in the global transcriptional profile between the two groups (**Figure 4B**). These results suggest that the duration of COVID-19 symptoms minimally impacts the resultant transcriptional profile of SARS-CoV-2 reactive CD4+ T cells in patients recently recovered from mild/moderate COVID-19.

**Figure 4.**
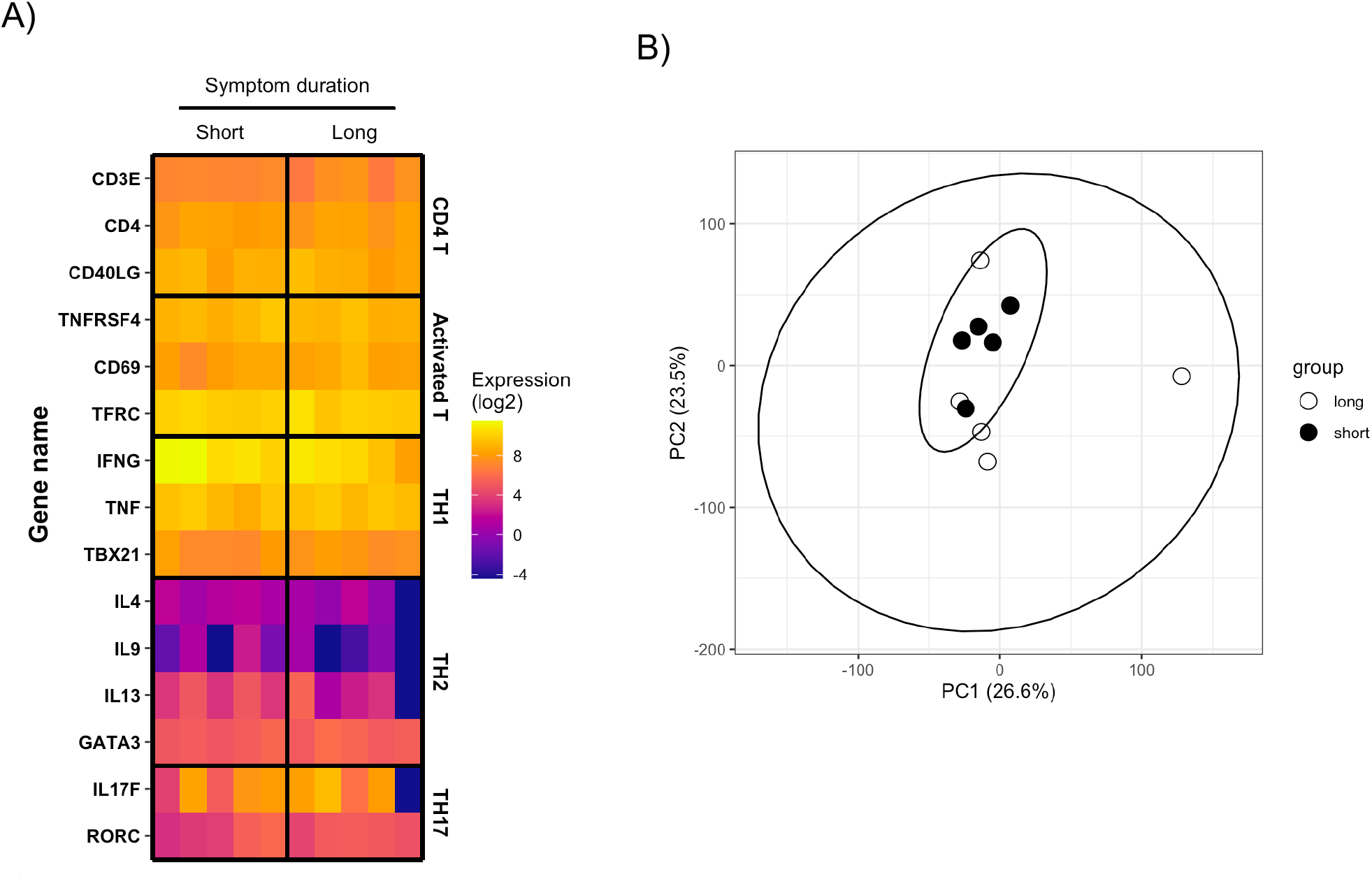
Transcriptional analysis of SARS-CoV-2 reactive CD4^+^ T cells. **A)** Heatmap display of normalized gene expression in sorted SARS-CoV-2 reactive CD4 T cells from total of 10 convalescent COVID-19 patients. Patients are separated by duration of self-reported COVID-19 symptoms. **B)** PCA analysis of total normalized gene expression data from sorted SARS-CoV-2 reactive CD4 T cells from total of 10 convalescent COVID-19 patients. Patients separated by duration of self-reported COVID-19 symptoms.

### Persistent COVID-19 symptoms do not correlate with seasonal coronavirus reactivity

Cellular immunity against SARS-CoV-2 antigens has been observed in PBMC samples collected prior to the emergence of the virus in December 2019 [19-21]. This has been primarily attributed to cross-reactive cellular immunity elicited by seasonal human coronaviruses such as 229E and NL63 which widely circulate and share some degree of antigen similarity with SARS-CoV-2 [19]. While the impact of pre-existing/cross-reactive cellular immunity on the clinical progression of SARS-CoV-2 infection remains unclear, we endeavored to determine if the presence of seasonal human coronavirus specific cellular immunity in convalescent COVID-19 patients correlated with the duration of self-reported symptoms, and if the magnitude of seasonal human coronavirus cellular immunity correlated with SARS-CoV-2 specific cellular immunity.

To this end, we utilized overlapping peptide pools spanning the Spike proteins of the human seasonal coronaviruses 229E and NL63 to stimulate PBMC from the same donors described above in a parallel IFN-γ ELISPOT assay. While the majority of subjects exhibited reactivity against the Spike protein from both 229E and NL63, persistent COVID-19 symptoms did not statistically impact magnitude of 229E (**Figure 5A**) or NL63 (**Figure 5B**) Spike protein reactivity as assessed by IFN-γ ELISPOT. While the magnitude of 229E and NL63 reactivity within a given subject correlated with each other, the magnitude of SARS-CoV-2 Spike protein reactivity observed in a given subject does not correlate with their reactivity to Spike from 229E or NL63 (**Figure 5C**), suggesting that these cellular populations may be distinct in convalescent COVID-19 patients. Interestingly, despite the lack of correlation between NL63/229E Spike reactivity and SARS-CoV-2 Spike reactivity, the presence of either NL63 or 229E Spike reactivity did correlate with reactivity against the SARS-CoV-2 N protein (**Figure 5C**). Finally, while we noted no significant association between the magnitude of SARS-CoV-2 specific cellular immunity and donor age in our study (**Supplemental Figure 4**), a significant negative correlation was observed between the total magnitude of 229E and NL63 reactivity and subject age in our dataset (**Figure 5D**).

**Figure 5.**
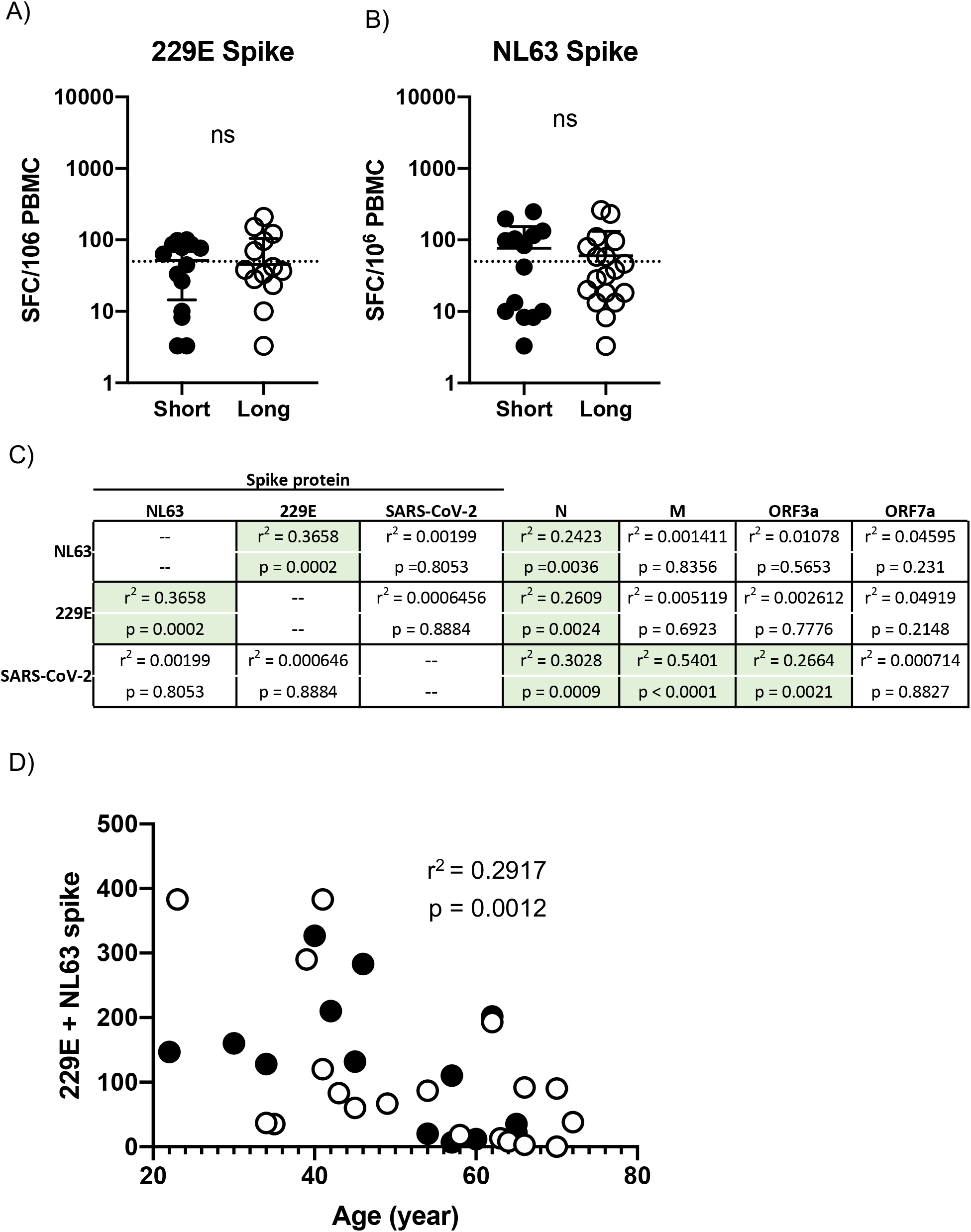
Assessment of seasonal human coronavirus cellular immunity in convalescent COVID-19 donors. **A)** Magnitude of 229E Spike protein specific cellular immunity in all study participants split by duration of self-report COVID-19 symptoms. **B)** Magnitude of NL63 Spike protein specific cellular immunity in all study participants split by duration of self-report COVID-19 symptoms. **C)** Correlation table assessing the relationship between NL63, 229E, and SARS-CoV-2 Spike protein reactivity in all subjects and the magnitude of reactivity against other human coronavirus antigens included in the study. r^2^ and p value calculated by 2-tailed Pearson Correlation test. **D)** Relationship between subject age and total 229E/NL63 Spike protein specific cellular immune response. Filled circles indicate subjects included in the “short” duration of symptoms group. Empty circles indicate individuals included in the “long” duration of symptoms group. r^2^ and p value calculated by 2-tailed Pearson Correlation test.

## DISCUSSION

In this study, we examined the relationship between the duration COVID-19 symptoms and the magnitude and functional profile of SARS-CoV-2 specific cellular immunity in individuals recently recovered from mild/moderate COVID-19. We observed that patients with prolonged COVID-19 symptoms overall exhibited similar levels of SARS-CoV-2 specific cellular immunity as individuals who rapidly resolved their symptoms. No defect was observed in the magnitude of the SARS-CoV-2 Spike specific CD4 and CD8 T cell response in individuals with prolonged COVID-19 symptoms when assessed using flow cytometry, and the transcription profile of SARS-CoV-2 specific CD4 T cells was observed not to be influenced by the duration of COVID-19 symptoms. Finally, while significant levels of cellular immunity against the seasonal human coronaviruses 229E and NL63 was observed in all convalescent COVID-19 patients analyzed in the study, the magnitude of this immune response did not correlate with the duration of COVID-19 symptoms. These data suggest that prolonged symptomatic COVID-19 does not significantly impact the development of SARS-CoV-2 specific cellular immunity in patients with mild/moderate disease.

The development of SARS-CoV-2 specific cellular immunity has been ubiquitously observed following the resolution of COVID-19 symptoms and may be a more sensitive immunologic indication of SARS-CoV-2 infection than conventional seroconversion [22, 23]. Indeed, higher levels of both cellular and humoral immunity have been observed in patients after the resolution of severe COVID-19 than following mild or asymptomatic infections [24]. However, the quality of the cellular immune response generated after severe COVID-19 is of uncertain quality, as severe COVID-19 is associated with severe T cell dysregulation, exhaustion, and inflammatory cytokine production [25, 26]. As we did not observe any appreciable deficit in either the quantity or the quality of the SARS-CoV-2 specific cellular immune profile in the individuals analyzed in our study, we feel that it is reasonable to hypothesize that the immunologic mechanisms underpinning severe COVID and persistent mild COVID-19 are distinct and may differentially impact the development of virus-specific cellular memory.

While it was not unexpected to observe cellular reactivity against coronaviruses other than SARS-CoV-2 in the convalescent COVID-19 patients analyzed in our study, the lack of correlation between the magnitude of SARS-CoV-2 Spike protein reactivity and reactivity against Spike from the seasonal coronaviruses 229E and NL63 was unexpected. Preexisting cellular immunity against common seasonal human coronaviruses – such as 229E, NL63, OC43, and HKU1-has been highlighted as the most likely explanation for the relativity high frequency of individuals with SARS-CoV-2 specific cellular immunity prior to the appearance of the virus [19-21]. Additionally, it is notable that the level of 229E and NL63 Spike reactivity did correlate with the amount of SARS-CoV-2 N reactivity in convalescent COVID-19 patients analyzed in our study. This result is consistent with other previously published reports that suggest that coronavirus infections – including infection with seasonal betacoronaviruses and SARS-CoV - may preferentially result in durable cellular memory against the structural N protein that cross-reacts with SARS-CoV-2 [21]. In addition to providing insight into the mechanisms driving the development of SARS-CoV-2 specific cellular immunity, this observation may provide guidance as to which antigens may be most amenable in the development of a universal coronavirus vaccine.

## METHODS

### Study design

Convalescent COVID-19 patients were recruited for this study at the SUNY Upstate Medical University Clinical Research Unit starting in March 2020 under the SUNY Upstate Convalescent Plasma Donor Program [27]. This study was reviewed by the SUNY Upstate Medical University IRB, reviewed approved by the Western Institutional Review Board (IRB # 1587400), and performed under informed consent. All subjects were adults 18 year of age or older who had previously tested positive for SARS-CoV-2 and who were symptom-free for at least 14 days prior to enrollment. Information regarding the timing and duration of acute COVID-19 symptoms – such as fever, shortness of breath, sore throat, cough that impacted activity, and fatigue that impacted activity -were self-reported. Lingering symptoms such as loss of taste and smell, mild cough or tickle in the throat, or lingering fatigue that did not impact their daily activity were not considered part of the acute illness and therefore not included in the length of illness. For asymptomatic donors identified via contract tracing protocols the date of positive RT-PCR test was used for the start and stop date of symptoms. Samples were de-identified following collection, and researchers conducting assays were blinded to clinical data until final comparative analysis. PBMC were collected and processed using Vacutainer CPT Cell Preparation Tubes (BD) and stored in vapor phase liquid nitrogen prior to analysis.

### IFN-γ ELISPOT

Cryopreserved PBMC were thawed, washed twice, and placed in RPMI 1640 medium (Corning, 10040CM) supplemented with 10% heat-inactivated Fetal Calf Serum (Corning, 35-010-CV), L-glutamine (Lonza, 17-605E), and Penicillin/Streptomycin (Gibco, 15140-122). Cellular viability was assessed by trypan blue exclusion and cells were resuspended at a concentration of 5×10^6^/ml and rested overnight at 37°C. After resting, viable PBMC were washed, counted, and resuspended at a concentration of 1×10^6^/ml in complete cell culture media. 100 μl of this cell suspension was mixed with 100 μl of the individuals peptide pools listed in Supplemental Table 2 and diluted to a final concentration 1 μg/mL/peptide (DMSO concentration 0.5%) in complete cell culture media. This cell and peptide mixture was loaded onto a 96-well PVDF plate coated with anti-IFN-γ (3420-2HW-Plus, Mabtech) and cultured overnight. Controls for each donor included 0.5% DMSO alone (negative) and anti-CD3 (positive). After overnight incubation the ELISPOT plates were washed and stained with anti-IFN-γ-biotin followed by streptavidin-conjugated HRP (3420-2HW-Plus, Mabtech). Plates were developed using TMB substrate and read using a CTL-ImmunoSpot® S6 Analyzer (Cellular Technology Limited). All peptide pools were tested in duplicate and the adjusted mean was reported as the mean of the duplicate experimental wells after subtracting the mean value of the negative (DMSO only) control wells. Individuals were considered reactive to a peptide pool when the background-subtracted response was >50 Spot Forming Cells (SFC)/10^6^ PBMC. All data were normalized based on the number of cells plated per well and are presented herein as SFC/10^6^ PBMC.

**Table 2.** Patterns of SARS-CoV-2 reactivity in convalescent COVID-19 patients

| **Virus** | **Antigen** | **Short duration of symptoms (< 8 days)** | **Long duration of symptoms (>18 days)** |
| --- | --- | --- | --- |
| SARS-CoV-2 | Any | 85.7% (12/14) | 94.7% (18/19) |
| SARS-CoV-2 | Spike | 71.4% (10/14) | 89.5% (17/19) |
| SARS-CoV-2 | N | 85.7% (12/14) | 57.9% (11/19) |
| SARS-CoV-2 | M | 50% (7/14) | 68.4% (13/19) |
| SARS-CoV-2 | ORF3a | 7.1% (1/14) | 26.3% (5/19) |
| SARS-CoV-2 | ORF7a | 0% (0/14) | 5.2% (1/19) |
| 229E | Spike | 50% (7/14) | 26.3% (5/19) |
| NL63 | Spike | 50% (7/14) | 36.8% (7/19) |

### Flow Cytometry

Surface staining for flow cytometry analysis was performed at room temperature in PBS supplemented with 2% FBS. Aqua Live/Dead (ThermoFisher, L34957) was used to exclude dead cells in all experiments. Antibodies and dilutions used for flow cytometry analysis are listed in Supplementary Table 2. Flow cytometry analysis was performed on a BD FACSAria II instrument and analyzed using FlowJo v10.7 software (Treestar).

### Isolation and transcriptional analysis of SARS-CoV-2 reactive CD4^+^ T cells

Cryopreserved PBMC samples were thawed and resuspended in complete cell culture media at a concentration of 5□×□10^6^ cells/mL and stimulated with 0.5□μg/mL of a SARS-CoV-2 Spike protein peptide pool (Supplementary Table 2) for 18□hours at 37□°C. Spike-reactive CD4^+^ T cells were identified by expression of the activation markers CD134 and CD69 and isolated by flow cytometric sorting using BD FACSAria II instrument. Cells were sorted directly into 350 μl RLT+ buffer (Qiagen, 1053393) supplemented with 1% 2-ME and RNA isolated using a RNeasy Micro spin column (Qiagen, 74004). cDNA was generated using a SMART-Seq HT Kit (TaKaRa, 634455), and the final Illumina-compatible DNA sequencing libraries were prepared using an Illumina Nextera XT DNA Library Preparation kit. RNA, cDNA, and DNA during the library preparation process were quantified using Agilent Bioanalyzer, and final libraries were sequenced using a 75 cycle High Output NextSeq 500/550 v2.5 reagent kit at the SUNY Upstate Molecular Analysis Core. Raw reads from FASTQ files were mapped to the human reference transcriptome (Ensembl, Home sapiens, GRCh38) using Kallisto [28] version 0.46.2. Transcript-level counts and abundance data were imported and summarized in R (version 4.0.2) using the TxImport package [29] and TMM normalized using the package EdgeR [30, 31]. Differential gene expression analysis performed using linear modeling and Bayesian statistics in the R package Limma [32].

### Statistical analysis

Statistical analyses were performed using GraphPad Prism v8 Software (GraphPad Software, La Jolla, CA). A *p*-value <0.05 was considered significant.

## Data Availability

The authors declare that all data supporting the findings of this study are available within this article or from the corresponding author upon reasonable request. RNAseq gene expression data have been deposited in the Gene Expression Omnibus database (GSE165373).

## AUTHOR CONTRIBUTIONS

HSF: Investigation, ADW: Investigation, KR: Investigation, Project administration, HF: Methodology, JRC: Methodology, SJT: Conceptualization, Project administration, Supervision, Methodology, TPE: Conceptualization, Project administration, Supervision, Methodology, ATW: Conceptualization, Data curation, Formal analysis, Funding acquisition, Investigation, Methodology, Visualization, Writing – original draft, Writing – review & editing

## ACKNOWLEDGEMENTS

We wish to thank all the study participants for making this work possible, Mark Abbott and Kristen Baxter for critical laboratory support, Lisa Phelps for support with the SUNY Upstate Medical University flow cytometry core, Karen Gentile for support with the SUNY Upstate Molecular Analysis Core, and Kristen Newell and Joel Wilmore for helpful discussion. This work was internally funded by the State University of New York. Material has been reviewed by the Walter Reed Army Institute of Research. There is no objection to its presentation and/or publication. The opinions or assertions contained herein are the private views of the author, and are not to be construed as official, or as reflecting true views of the Department of the Army or the Department of Defense. The investigators have adhered to the policies for protection of human subjects as prescribed in AR 70-25

---

**Supplemental Figure 1.**
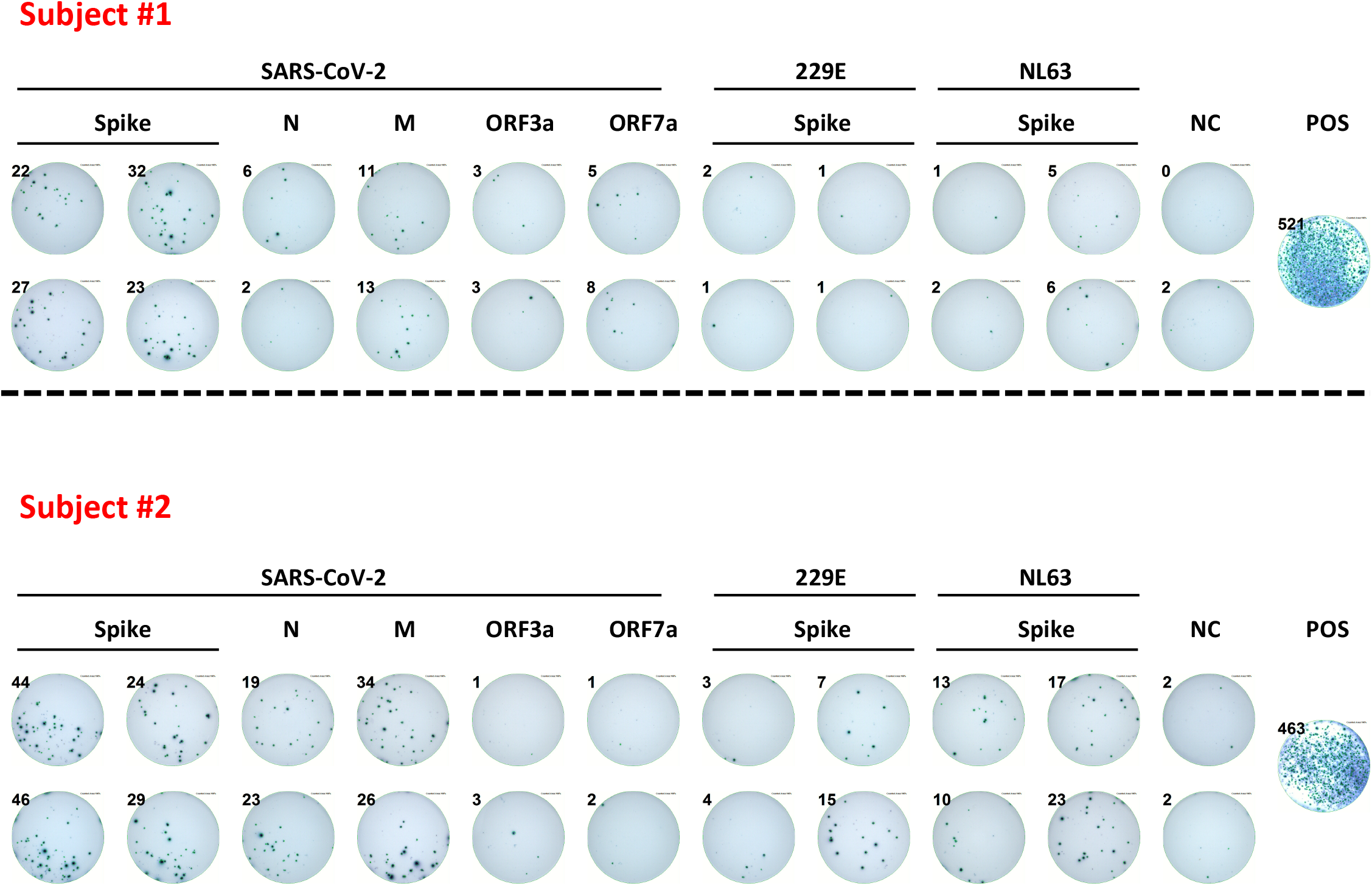
Representative IFN-γ ELISPOT plate images

**Supplemental Figure 2.**
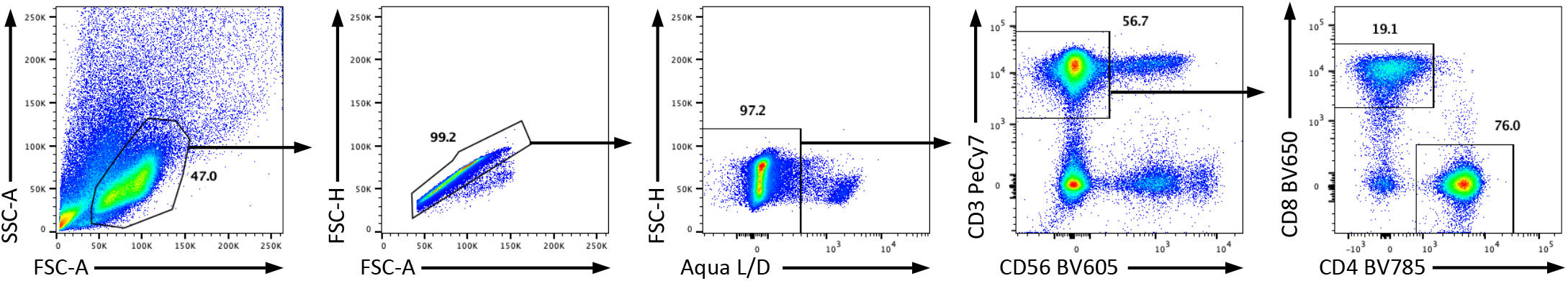
Gating scheme for activated CD4 and CD8 T cell isolation

**Supplemental Figure 3.**
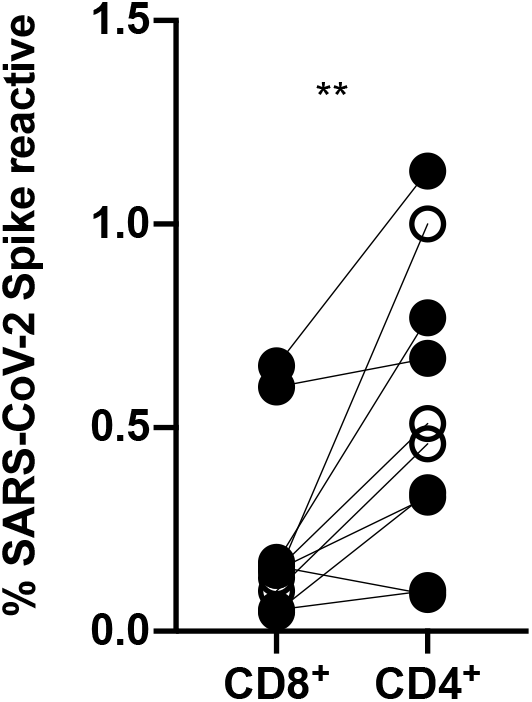
Background-subtracted frequency of SARS-CoV-2 Spike reactive CD8+ and CD4+ T cells in 10 select convalescent COVID-19 patients. ^**^ p < 0.01 two-tailed paired T test.

**Supplemental Figure 4.**
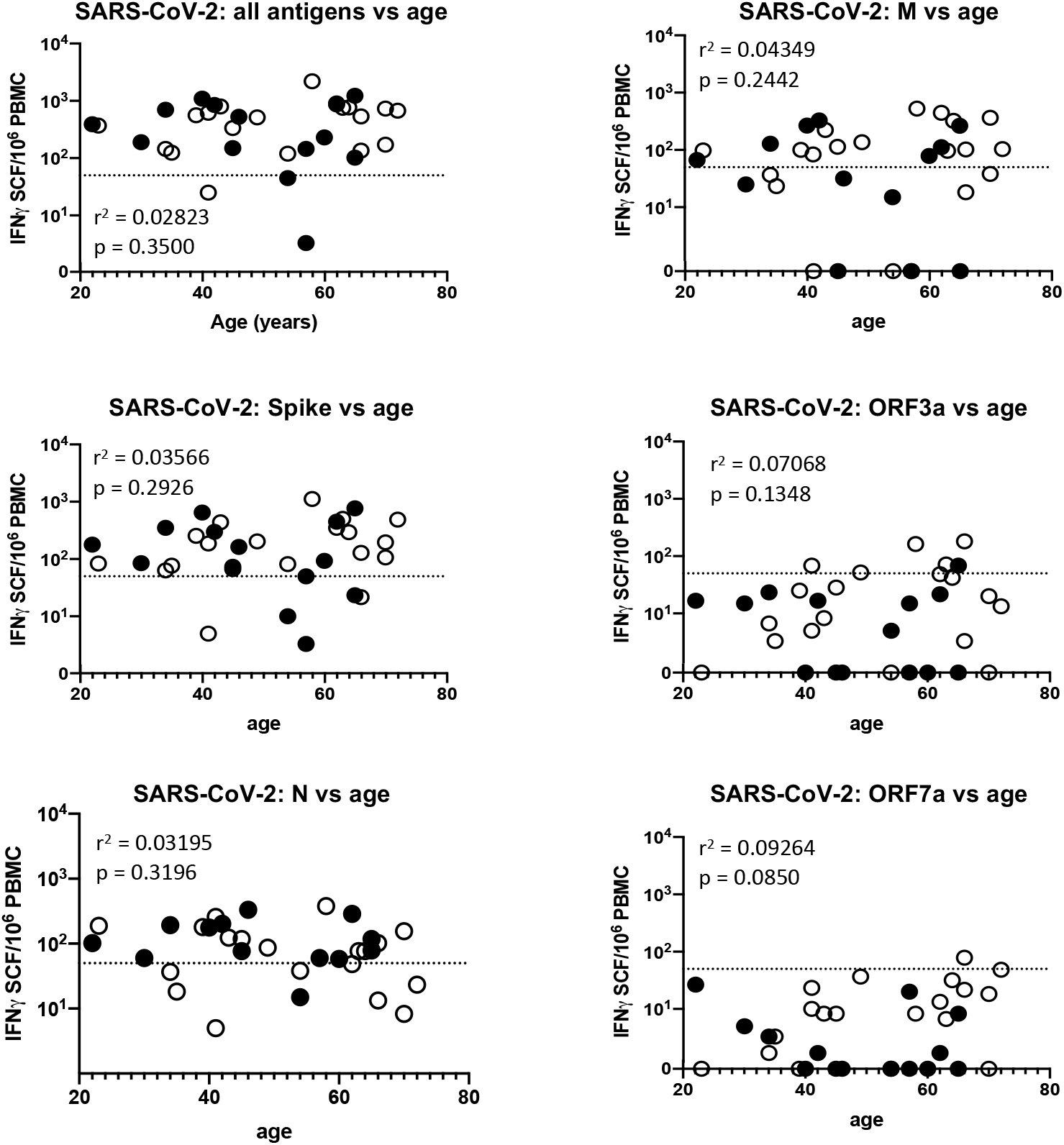
Relationship between age and SARS-CoV-2 antigen reactivity as assessed by IFN-g ELISPOT. Pearson correlation calculations were used to determine r2 and two-tailed p values. Dashed line indicates the 50 IFN-γ SFC/10^6^ PBMC threshold for positivity

