## Supplemental tables for "Persistent COVID-19 symptoms minimally impact the development of SARS-CoV-2 specific cellular immunity"

**Supplemental Table 1.** Peptide pools used for IFN-g ELISPOT analysis

| **Virus** | **Antigen** | **Company** | **Product #** |
| --- | --- | --- | --- |
| SARS-CoV-2 | Spike glycoprotein | JPT Peptide Technologies | PM-WCPV-S |
| SARS-CoV-2 | Nucleoprotein peptide pool | JPT Peptide Technologies | PM-WCPV-NCAP |
| SARS-CoV-2 | Membrane peptide pool | JPT Peptide Technologies | PM-WCPV-VME |
| SARS-CoV-2 | ORF3A peptide pool | JPT Peptide Technologies | PM-WCPV-AP3A |
| SARS-CoV-2 | ORF7A peptide pool | JPT Peptide Technologies | PM-WCPV-NS7A |
| HCoV-229E | Spike glycoprotein | JPT Peptide Technologies | PM-229E-S-1 |
| HCoV-NL63 | Spike glycoprotein | JPT Peptide Technologies | PM-NL63-S-1 |

**Supplemental Table 2.** Antibody used for flow cytometry

| **Antibody** | **Clone** | **Manufacturer** | **Catalog #** | **Lot #** | **Dilution used** |
| --- | --- | --- | --- | --- | --- |
| CD25 PE | BC69 | Biolegend | 302606 | B296102 | 1:160 |
| CD3 PeCy7 | HIT3a | Biolegend | 300316 | B293468 | 1:320 |
| CD69 AF647 | FN50 | Biolegend | 310918 | B275384 | 1:160 |
| CD134 FITC | Act36 | Biolegend | 350006 | B296280 | 1:160 |
| CD56 BV605 | 5.1H11 | Biolegend | 362537 | B320741 | 1:160 |
| CD8 BV650 | RPA-T8 | Biolegend | 301041 | B275821 | 1:320 |
| CD4 BV785 | RPA-T4 | Biolegend | 300553 | B298451 | 1:160 |

**Supplemental Table 3.** RNAseq metrics

| **Sample ID** | **Duration of symptoms (days)** | **Number of sorted cells** | **RNA recovery** | **# aligned reads** |
| --- | --- | --- | --- | --- |
| Short #1 | 2 | 3,179 | 2,370pg | 45.6M |
| Short #2 | 5 | 4,536 | 3,255pg | 47.0M |
| Short #3 | 3 | 648 | 496pg | 43.6M |
| Short #4 | 5 | 1,318 | 1,170pg | 48.1M |
| Short #5 | 7 | 4,076 | 2,325pg | 46.0M |
| Long #1 | 19 | 2,214 | 1,095pg | 45.1M |
| Long #2 | 18 | 1,586 | 765pg | 45.2M |
| Long #3 | 33 | 4,497 | 2,025pg | 49.6M |
| Long #4 | 61 | 1,061 | 1,155pg | 46.6M |
| Long #5 | 37 | 106 | 690pg | 45.0M |
